# Machine learning applied to atopic dermatitis transcriptome reveals distinct therapy-dependent modification of the keratinocyte immunophenotype

**DOI:** 10.1101/2019.12.14.19014977

**Authors:** K. Clayton, A. Vallejo, S. Sirvent, J. Davies, G. Porter, F. Lim, M.R. Ardern-Jones, M.E. Polak

**Affiliations:** Clinical and Experimental Sciences, Sir Henry Wellcome Laboratories, Faculty of Medicine, University of Southampton, Southampton, United Kingdom; Institute for Life Sciences, University of Southampton, United Kingdom; Department of Dermatology, University Hospitals Southampton NHS Foundation Trust, United Kingdom; Unilever, Colworth Science Park, Sharnbrook, Bedford, United Kingdom

## Abstract

**Background:** Atopic dermatitis (AD) arises from a complex interaction between an impaired epidermal barrier, environmental exposures, and the infiltration of Th1/Th2/Th17/Th22 T cells. Transcriptomic analysis has advanced understanding of gene expression in cells and tissues. However, molecular quantitation of cytokine transcripts does not predict the importance of a specific pathway in AD or cellular responses to different inflammatory stimuli.

**Objective:** To understand changes in keratinocyte transcriptomic programmes in human cutaneous disease during development of inflammation and in response to treatment.

**Methods:** We performed *in silico* deconvolution of the whole-skin transcriptome. Using co-expression clustering and machine learning tools, we resolved the gene expression of bulk skin (n=7 datasets, n=406 samples), firstly, into unsupervised keratinocyte immune response phenotypes and, secondly, into 19 cutaneous cell signatures of purified populations from publicly available datasets.

**Results:** We identify three unique transcriptomic programmes in keratinocytes, KC1, KC2, KC17, characteristic to immune signalling from disease-associated helper T cells. We cross-validate those signatures across different skin inflammatory conditions and disease stages and demonstrate that the keratinocyte response during treatment is therapy dependent. Broad spectrum treatment with ciclosporin ameliorated the KC17 response in AD lesions to a non-lesional immunophenotype, without altering KC2. Conversely, the specific anti-Th2 therapy, dupilumab, reversed the KC2 immunophenotype.

**Conclusion:** Our analysis of transcriptomic signatures in cutaneous disease biopsies reveals the complexity of keratinocyte programming in skin inflammation and suggests that the perturbation of a single axis of immune signal alone may be insufficient to resolve keratinocyte immunophenotype abnormalities.

## Introduction

Atopic dermatitis (AD) arises from a complex interaction between impaired epidermal barrier and environmental exposures to allergens and irritants, resulting in aberrantly activated infiltrating immune cells. Much interest has focused on the immune cells infiltrating AD skin, which mediate the disease. In particular, dense infiltration of activated Th2/Th22 CD4+ T cells has been observed as an early feature of AD exacerbations, especially in acute lesions^1^. These are identified also in non-lesional skin of AD sufferers suggesting a systemic immunodysregulation^2,3^. This implies that type 2 cytokines play a major role in disease pathogenesis and clinical research has shown the impressive efficacy in AD treatment of a monoclonal antibody therapy targeting IL4Ra which blocks IL-4 and IL13 signalling^4^. However, studies by Gittler *et al*. first demonstrated that the Th1/Th17 axis is also prominent in chronic AD lesions and correlates with the magnitude of the Th2 signals^5^. Whilst various different T cell pathways have been targeted in clinical trials of AD, the functional effects of the inflammatory pathways on skin keratinocytes have largely been ignored^6,7^.

Alongside the immune skin infiltrate, spongiosis and keratinocyte hyperplasia are the cardinal features of epidermal changes in AD. In addition to the gene mutation mediated reduction of filaggrin expression, type 2 inflammation also reduces keratinocyte filaggrin expression thereby further damaging the skin barrier^8,9^. Importantly, beyond their role in maintaining the physical barrier of the skin, keratinocytes also act as innate immune sentinels, and express pattern recognition receptors, ligation of which regulates keratinocyte synthesis of cytokines, anti-microbial peptides (AMPs) and antigen presentation to immune cells^10–16^.

The role of IL-17 and IL22 cytokines in regulating antimicrobial peptides such as s100 proteins and β-defensins is well established^17–22^ and the importance of these pathways in psoriasis has been validated by clinical demonstration of effectiveness of inhibitory monoclonal antibody therapy^23,24^; their precise function in AD is less clear. To study the key pathways driving AD, where targeted intervention may prove most fruitful, direct quantitation of the immune signals (e.g. cytokines) can be undertaken. However, as molecular quantitation of the cytokine transcripts does not predict the importance of a specific pathway in AD, it is necessary to study the outcome of the epidermal responses to different inflammatory stimuli to properly define their role. Thus, to characterise the keratinocyte immunophenotype it is necessary to be able to investigate skin transcriptome to a cellular resolution. Single-cell analysis can offer an approach to this question but is limited by the technical challenge of achieving adequate encapsulation of enough cells of interest with minimal transcriptomic disturbance. Here we show that it is possible to employ machine learning to resolve the keratinocyte transcriptomic signal from the non-keratinocyte skin transcriptome, revealing important insights to the pathogenesis of AD.

## Methods

### Microarray data analysis

Microarray datasets were obtained from the Gene Expression Omnibus (GEO, NCBI) and were analysed from raw data in R and normalised according to platform specifics. For Affymetrix microarray platforms, classic microarray quality control was performed using the Bioconductor *arrayQualityMetrics* tool and were normalised to obtain expression values by GCRMA methods within the Affymetrix package. Illumina platforms and other technologies were quantile normalised using the *lumi* or *limma* Bioconductor packages.

### Unsupervised network clustering

The inflammatory skin disease datasets GSE32924, GSE36842 and GSE34248 were processed as above and subsequently merged and batch corrected using the *COMBAT* tool within the SVA Bioconductor package. Differential gene expression analysis between healthy controls and lesional skin, within disease lesional and non-lesional, and across disease lesional comparisons was conducted using a filtering of Benjamini-Hochberg adjusted p-value >0.05, log(2)-fold difference x1 using the LIMMA Bioconductor package. The expression values of 4620 probset-IDs, corresponding to 3066 unique genes were input into MIRU (now, GraphiaPro) for network analysis^25,26^. A transcript-to-transcript correlation matrix using Pearson correlation coefficient of *r* ≥0.7 was created. The resulting network graph was then clustered into groups of genes using the MCL algorithm at an inflation value of 3.1 and minimum cluster size of 10 genes, giving 50 clusters. The gene list for each cluster was interrogated for gene ontology using the web-based analysis tool ToppFun within the ToppGene suite^27^. The REVIGO online tool was used to provide a single biological process term for each cell-based cluster, selecting that with both the lowest B.H. p-value and a term dispensability of zero.

### Reference cutaneous cell populations datasets

To curate cell profiles for input as CIBERSORT reference signatures, we collated seven datasets from GEO: GSE36287 (keratinocytes stimulated with IFNα, IFNγ, IL-13, IL-17A, IL-4, TNFα, and unstimulated control), GSE7216 (keratinocytes stimulated with IL-1β, IL-22, IL-26, keratinocyte growth factor (KGF)), GSE34308 (dermal fibroblasts), GSE74158 (skin resident CD4, CD8 and regulatory T cells), GSE4570 (melanocytes), GSE49475 (activated Langerhans’ cells, CD11c^+^ dermal dendritic cells), and GSE23618 (steady-state Langerhans’ cells). Datasets were normalised separately, and gene expression of sample replicates averaged by mean before combining into a single file for upload to CIBERSORT as a signature genes matrix. Gene replicates are discounted by CIBERSORT in favour of the gene with the highest mean expression across the samples.

### Bulk skin datasets

Datasets of skin biopsies from inflammatory skin diseases were obtained from GEO. Four AD datasets and two psoriasis datasets were analysed. GSE32924 is a dataset of paired chronic lesional (AD-ChL, n=11), non-lesional (AD-Non, n=11) atopic dermatitis samples, and healthy controls (HH, n=8). GSE36842 is a dataset of acute lesional (AD-AcL, n= 7), chronic lesional (AD-ChL, n=7), non-lesional (AD-Non, n=7) skin from atopic dermatitis patients and healthy controls (HH, n=6). GSE58558 is a longitudinal study of ciclosporin treatment of atopic dermatitis patients with biopsies taken at baseline, 2 and 12 weeks of treatment of both chronic lesions (CAL: baseline, n=16; 2-weeks, n=17; 12-weeks, n=17) and non-lesional skins (ANL: baseline, n=16; 2-weeks, n=16; 12-weeks, n=16). GSE130588 is a longitudinal study of dupilumab or placebo treatment of atopic dermatitis patients with biopsies taken at baseline, 4 and 16 weeks of treatment of both chronic lesions (AD-ChL: baseline-dupilumab, n=26; baseline-placebo, n=25; 4-weeks-dupilumab, n=24; 4-weeks-placebo, n=20; 16-weeks-dupilumab, n=16; 16-weeks-placebo, n=16) and non-lesional skins (AD-Non: baseline-dupilumab, n=22; baseline-placebo, n=20; 16-weeks-dupilumab, n=15; 16-weeks-placebo, n=7) and healthy controls (HH, n=20). GSE34248 is a dataset of paired lesional (Ps-Les, n=14) and non-lesional (Ps-Non, n=14) psoriatic samples. GSE11903 is a longitudinal study of etanercept treatment of psoriasis patients with biopsies taken at baseline for lesional and non-lesional skin (Ps-Les, n=11; Ps-Non, n=11) and biopsies from lesional skin at 1- (n=11), 2- (n=11), 4- (n=10), and 12 weeks (n=11) of treatment

### Running CIBERSORT

Datasets of bulk skin samples were deconvoluted against the reference signature sets using the online version of the CIBERSORT^28^ algorithm. Reference signature files were provided as the *signature gene file*, while normalised expression data of bulk samples were provided as the *mixture file*. All run settings were kept at default. Output provided by CIBERSORT was downloaded as a .txt file, where relative abundance of each cellular signature was normalised as a percent of sample.

## Results

### Gene co-expression analysis of inflammatory skin disease genes reveals immune and keratinocyte involvement

Firstly, we set out to examine the transcriptomic signals from skin biopsies of lesional, regardless of chronicity, and non-lesional AD and psoriasis (AD-Les, AD-Non, Ps-Les, Ps-Non, respectively). In line with previous reports, unsupervised differential expression analysis identified 4620 genes^3,29^. Transcript-to-transcript clustering (GraphiaPro, Pearson r >0.7, MCL=3.1, >10 clustered genes) identified 50 clusters (Figure 1a). Annotation revealed three clusters (11, 14 and 28) encoding immune-related processes such as lymphocyte activation, interferon and cytokine signalling (Figure 1f-h; Supplementary table 1). The relative expression pattern across these clusters showed similar changes in lesions, regardless of disease, and the least expression in healthy skin. Interestingly, AD non-lesional skin showed a prominent defence response (cluster 28), suggesting a subclinical immune alteration in non-lesional AD skin as compared to healthy skin^3,30,31^.

**Figure 1 legend.**
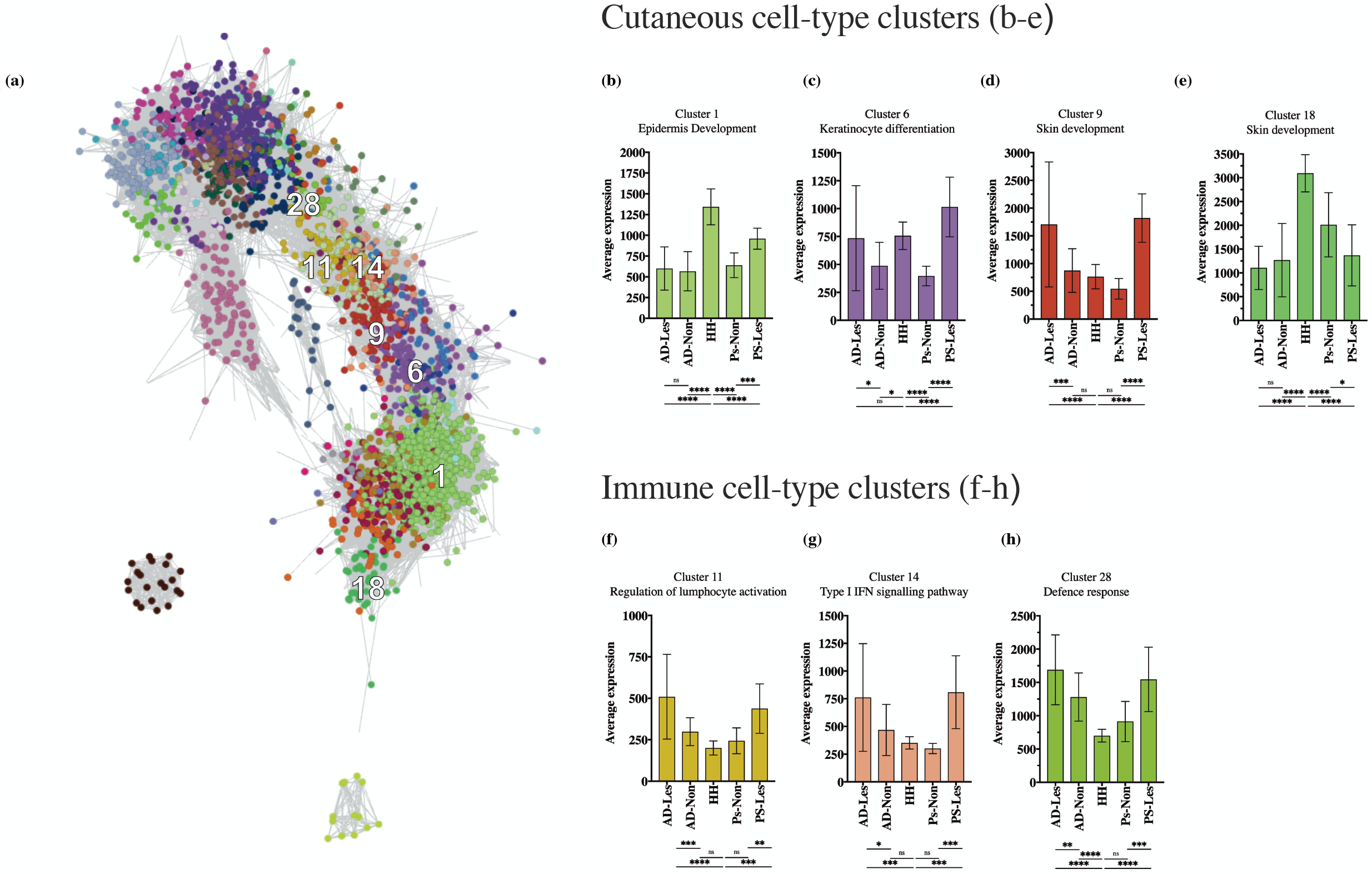
Unsupervised co-expression analysis of whole skin from healthy, atopic and psoriatic skin. **(a)** Transcript-to-transcript clustering of 4620 differentially expressed genes of atopic lesional (AD-Les), atopic non-lesional (AN-Non), psoriatic lesional (Ps-Les) and psoriatic non-lesional (Ps-Non) compared to healthy control (HH) (FDR p-value <0.05, log fold-change <-2/>2). Gene to gene co-expression correlation of >0.7 Pearson were retained for Markov clustering using an inflation value of 3.1. **(b-h)** The top 50 clusters were annotated for biological process, of which seven clusters were identified as cell-based. Average expression of the all the genes in cluster shown per phenotype. ANOVA Sidak’s multiple test bars below: p>0.05,ns; p<0.05, *; p<0.01, **; p<0.001, ***; p<0.0001, ****. Error bars show mean± SD.

Strikingly, clusters 1, 6, 9 and 18 (Figure 1b-e; Supplementary table 1) were enriched in biological processes characteristic for keratinocytes (KCs). Genes in clusters 1 and 18 were most highly expressed in healthy tissue and represented processes of epidermis development and skin development, respectively, suggesting aberrant regulation of these processes in inflammatory skin disease. In contrast, genes in cluster 9 showed strong correlation with expression of immune mediated inflammation genes which suggests that the association may be causal. Cluster-to-cluster gene expression by tissue showed high correlation (Pearson r-square >0.7 (data not shown)). It is notable that genes in cluster 9 include *KRT6A/B, KRT16*, and the *S100* and *SERPIN* encoding proteins which are known to be involved in epidermal perturbation from inflammation and hyper-proliferative barrier breach^2,32,33^, indicating that the cross-talk between immune inflammation and keratinocyte function is important for lesion pathogenesis.

### Machine learning resolution of whole skin samples into constituent cellular profiles

To resolve from bulk expression data the transcriptomic signatures of keratinocyte responses we utilised a machine learning approach. We trained an algorithm (CIBERSORT^28,34^) to resolve gene expression profiles of purified cellular populations and tested this on whole tissue transcriptomic data from split skin (epidermis and dermis) to identify the relative proportion of cells. We then utilised laser captured dermal and epidermal regions from both healthy and AD skin^35^ to demonstrate that the algorithm could reliably separate relative proportions of the keratinocyte and fibroblast composition of whole skin in uninflamed and inflamed settings (Figure 2a, b). We expanded this approach by utilising a training set of transcriptomes for melanocytes, resident regulatory, CD4 and CD8 T cells, steady-state and activated Langerhans cells, and CD11c^+^ dermal dendritic cells to increase the resolution of the cellular skin components (Figure 2c).

**Figure 2 legend.**
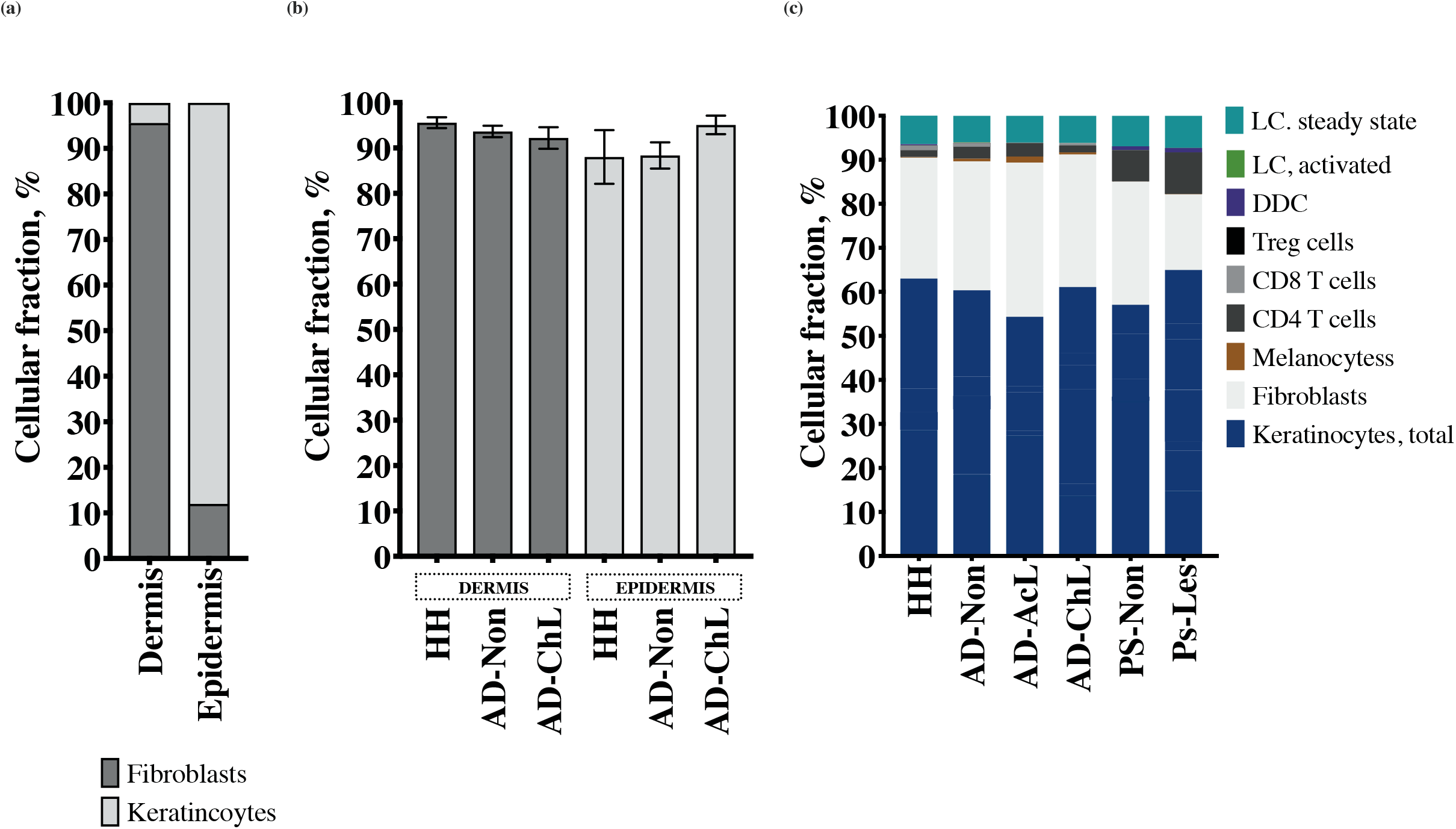
Machine learning resolution of whole skin samples into constituent cellular profiles. **(a)** Identification of key cellular signatures (fibroblasts: dark grey and keratinocytes: light grey) in microdissected samples from dermis and epidermis by machine learning (healthy dermis, n=6; healthy epidermis, n=10). **(b)** Inflammation/disease status (HH, healthy; AD-Non, AD non-lesional sample; AD-ChL, AD chronic lesion sample) of dermis or epidermis does not affect the correct deconvolution of cellular components within the bulk disease tissue (dermis: HH, n=6; AD-Non, n=5; AD-ChL, n=5. Epidermis: HH, n=10; AD-Non, n=5; AD-ChL, n=5). (**a,b)** (default CIBERSORT settings: 100 permutations, kappa = 999, q-value = 0.3, number of barcode genes 50-150. Training signatures: GSE36287, unstimulated keratinocytes; GSE34308, skin fibroblasts, Supplementary table E2) **(c)** Whole skin from healthy controls and AD patients (non-lesional, AD-Non; acute and chronic lesional, AD-AcL and AD-ChL, respectively) resolved into relative fractions of cutaneous cell populations of 9 transcriptomic signatures; keratinocytes; fibroblasts; melanocytes; CD4+, CD8+ and regulatory T cells; dermal dendritic cells; and, steady-state and activated Langerhans cells). The mean percentage of each of the signatures is shown relative to the remaining signatures. Healthy controls (n=14), and AD patients (non-lesional, n=18; acute lesional, n=7; chronic lesional, n=18). (default CIBERSORT settings. Training signatures: Supplementary table E3).

### *In silico* sorting reveals prevalence of KC2 and progression of KC17 immunophenotypes during the course of the inflammation in AD

To identify sub-populations of keratinocytes showing a molecular response to a specific inflammatory cytokine, we further trained the algorithm to resolve keratinocytes responding to IFN-α and IFN-γ (KC1), IL-4 and IL-13 (KC2), IL-17A (KC17), KGF, IL-26, IL-22, TNF, IL-1β, as well as resting (Figure 3b-e, Supplementary figure S1). These were tracked across 14 healthy (HH), 18 AD non-lesional samples (AD-Non), seven AD acute lesional samples (AD-AcL) and 18 AD chronic lesional samples (AD-ChL) (GSE32924 and GSE36842) and 14 psoriasis non-lesional samples (Ps-Non) and 14 in psoriasis lesional (Ps-Les) to investigate disease-related shift in the transcriptomic programme of these cells (Figure 3b-e). As expected, in AD, a strong KC2 signal was clearly detectable, showing that keratinocytes in chronic lesions were significantly responding to type 2 cytokines in comparison to healthy skin (ANOVA, p<0.05) (Figure 3c). Interestingly, this signal was equally strong in non-lesional skin suggesting that this dysregulation may be systemic in AD.

**Figure 3 legend.**
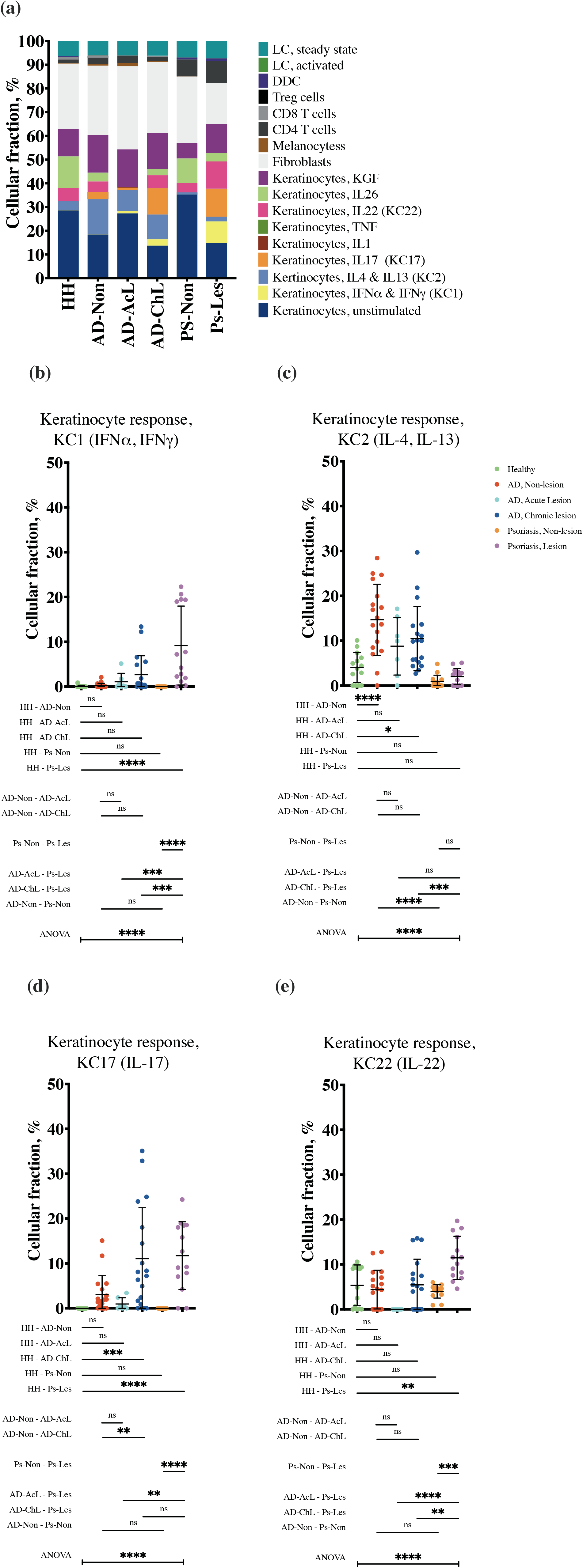
Resolution of whole skin samples into constituent cellular profiles using machine learning. **(a)** Deconvolution of keratinocyte signature into profiles representing keratinocyte immunophenotypes across the six skin conditions: healthy (HH), AD non-lesional (AD-Non), AD acute lesional (AD-AcL), AD chronic lesional (AD-ChL), psoriasis non-lesional (Ps-Non), psoriasis lesional (Ps-Les). (Default CIBERSORT settings, training signatures: keratinocytes (unstimulated, stimulated with IFNα, IFNγ, IL-17, IL-1β, TNF, IL-22, IL26, KGF); fibroblasts; melanocytes; CD4+, CD8+ and regulatory T cells; dermal dendritic cells; and, steady-state and activated Langerhans cells here, Supplementary table E4) **(b-e)** Individual keratinocyte immunophenotypes; **(b)** KC1 (response to IFNα, IFNγ); **(c)** KC2 (response to IL-4, IL-13); **(d)** KC17 (response to IL-17), **(e)** KC22 (response to IL-22); remaining KC fractions shown in Supplementary figure E1. Healthy patients (n=14, green), AD non-lesion patients (n=18, red), AD acute lesion patients (n=7, cyan), AD chronic lesion patients (n=18, blue), psoriasis non-lesion patients (n=14, orange), and psoriasis lesion patients (n=14, magenta). ANOVA Sidak’s multiple test bars below: p>0.05,ns; p<0.05, *; p<0.01, **; p<0.001, ***; p<0.0001, ****. Error bars show mean± SD.

A keratinocyte interferon programme, KC1, was prominent in lesional and absent from non-lesional psoriatic samples (ANOVA, p<0.0001) (Figure 3b). KC1 was also found in chronic lesions from AD samples, showing a trend of increase from non-lesional and acute lesional stages (Figure 3b), inferring the complex Th2/Th17/Th1 inflammation experienced by the epidermis of chronic AD lesions also alters the transcriptomic programme of keratinocytes. As expected, the KC17 signal was significantly elevated in psoriatic lesions compared to heathy controls and paired non-lesion samples (ANOVA, p<0.0001) (Figure 3d). In contrast to AD, the Th17 signal was absent in both non-lesional psoriatic skin and skin of healthy controls. Compared to healthy controls, AD keratinocytes showed an IL-17 sensing signal in lesional as well as non-lesional skin, but this was only significant for chronic lesions (ANOVA p<0.0001) (Figure 3d). KC17 appeared to dominate in chronic AD lesions, suggesting an evolution in Th17 pathway over time.

### Treatment specific modification of keratinocyte immunophenotype signature

Microarray data from psoriasis patients treated with etanercept, showed longitudinal abrogation of KC17 (p<0.0001) underscoring the role of this pathway in psoriasis pathogenesis (Figure 4b). The immunophenotypic signature of response to AD treatment was more nuanced. Despite efficacy and improvement in disease severity scores across the cohort (SCORAD50 improvement), ciclosporin treatment did not reduce the KC2 fraction in either non-lesional or lesional samples (Figure 4c). However, the KC17 fraction in lesional keratinocytes was significantly reduced by ciclosporin (p=0.02) which evolved to show no significant difference from non-lesional skin (p>0.05) (Figure 4d).

**Figure 4 legend.**
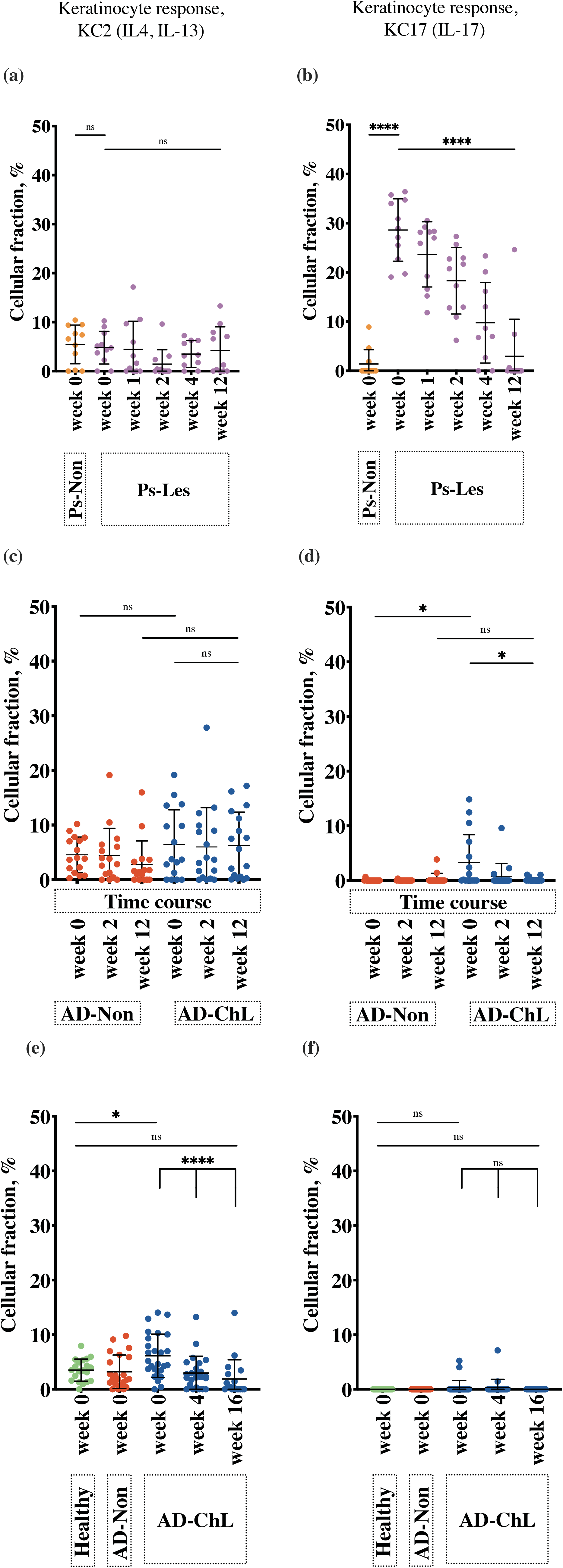
Response of disease-related keratinocyte fractions, KC2 and KC17, to treatment of psoriasis and AD. **(a, b)** KC2 **(a)** and KC17 **(b)** in whole skin from psoriasis patients undergoing etanercept treatment (non-lesional (week 0 only, n=11, orange), and lesional (weeks 0, 1, 2, and 12, n=11; week 4, n=10, magenta) [GSE11903 (Default CIBERSORT settings, training signatures: as Figure 3, Supplementary table E4)]. **(c, d)** KC2 **(c)** and KC17 **(d)** during a time course of ciclosporin treatment at baseline (AD-Non (red), n=16; AD-ChL (blue), n=16), mid-treatment (2 weeks) (ANL, n=17; CAL, n=16), and end of treatment at 12 weeks (ANL, n=17; CAL, n=16). [GSE58558 (Default CIBERSORT settings, training signatures: as Figure 3, Supplementary table E4)] **(e, f)** KC2 **(e)** and KC17 **(f)** fractions in healthy (green), AD non-lesional (red) and AD lesional (blue) AD samples during a time course of dupilumab treatment at baseline (HH, n=20; AD-Non, n=42; AD-ChL, n=51), mid-treatment (4 weeks) (AD-ChL, dupilumab-treated n=24), and end of treatment (16 weeks) (AD-ChL, dupilumab-treated n=18). [GSE130588 (Default CIBERSORT settings, training signatures: as Figure 3, Supplementary table E4)].

To follow the effect of anti-Th2 treatment on keratinocyte transcriptomic responses, we used the GEO microarray dataset GSE130588, where bulk skin microarray data was obtained from AD patients treated with 400mg of dupilumab with biopsies taken at treatment initiation, 4-weeks and 16-weeks on the conclusion of treatment, along with a cohort healthy control. Biopsies were taken from the same lesional site and showed a clinical and transcriptomic improvement in clinical atopy with treatment in established, ongoing lesions. Interestingly, very few of the AD samples, including many of the lesional biopsies, showed a KC17 fraction. Dupilumab treatment demonstrated a dramatic early reduction in the KC2 fraction of lesions at 4 weeks compared to baseline (p=0.0007), which was sustained at 16 weeks (p=0.0002) (Figure 4e). At the start of treatment, AD lesions had an increased KC2 profile compared to healthy controls, which resolved to be comparable by the end of anti-IL-4/IL-13 treatment.

## Discussion

Despite our understanding of the role of inflammatory cells in AD, and identification of the different inflammatory signals evident in AD, it is surprising that little attention has been paid to characterising the keratinocyte response in detail. This has perhaps, in part, been due to technical challenges associated with addressing the question. Standard approaches to bioinformatic analysis of transcriptomic studies of AD^4^ employ statistical tools to identify differentially expressed genes in lesions versus non-lesions and can utilise gene set enrichment analysis based on the functional annotation of the differentially expressed transcripts to identify cellular processes which are more or less prominent in AD.

However, such a macro view of the AD transcriptome prevents characterisation of individual responses of the various cell-types comprising skin. Widely used approaches to look at individual cell populations include flow cytometry and immunohistochemistry. Despite the routine application of these methodologies, and valuable insights they can provide for a relatively narrow set of markers, they only inform analysis of cell phenotype in a relatively limited way, partly because both of these techniques require a monoclonal antibody label of which a limited panel can be applied to a single sample. Single cell sequencing from whole skin or from flow sorted populations would allow the investigator to undertake detailed characterisation of cell types in AD in a non-hypothesis driven manner. However, as yet, these approaches are limited by cost, and the relatively low number of sequenced cells as a proportion of the whole tissue sample may present a sampling bias. We sought to address cellular analysis within tissue by a different approach. Using machine learning on published datasets, we trained an algorithm to identify different skin cell types and then cellular responses to different inflammatory responses of interest. We validated this algorithm on transcriptomic studies of microdissected healthy skin and psoriasis which has a known immune pathway dependence, showing that our approach is a powerful method for investigating transcriptomic signatures in skin samples of complex disease.

Applying such optimised machine learning-based analysis to existing datasets of AD disease stages and during treatment has confirmed the constitutive atopic skin phenotype in AD patients. An altered transcriptomic programme in keratinocytes was evident in all samples (lesional and non-lesional) which reflected Th2 sensing by keratinocytes which we termed “KC2”, and similar findings have been reported by others^3,5^. Further, we show the immunophenotype shift characteristic of lesion progression modifies keratinocyte profiles to an IL-17 (KC17) and interferon (KC1) sensing phenotype. Thus, we could demonstrate that although acute AD lesions show a strong Th2 signal, and chronic lesions have Th1 and Th17 signals, the Th2-related processes is amplified rather than a switch away from a type 2 cytokine response in chronic lesions.

Effective treatment of psoriasis with ciclosporin showed a reversal of the dominant KC17 profile of lesional skin to that of unaffected skin. However, in AD, remarkably, and despite resolution of skin inflammation as measured by disease severity, ciclosporin did not modify the KC2 profile of lesional skin. Instead, AD disease remission with ciclosporin correlated with loss of the KC17 signal. In contrast to this, with dupilumab, striking loss of the KC2 signal associated with disease remission, whereas the KC17 response was not observed. This underscores the critical role of the type 2 cytokines in AD and might suggest a strong role for IL-17 pathway in AD pathogenesis; it may also reflect the complex immunophenotype of the disease and potential immune mediator redundancy.

Our analysis to computationally resolve keratinocyte sub-populations by their sensing of immune-related signals does not address AD as a disease driven by epidermal disruption or systemic immune abnormalities. Indeed, we see evidence of some individual variation at a keratinocyte level, particularly of KC2 and KC17 immunophenotypes. We postulate that various environmental factors such as commensal dysbiosis may contribute to individual variation in epidermal sensing by keratinocytes and may regulate the epidermal response, both modifying and modified by immune infiltrate signals. This theory would suggest that perturbation of the immune signal alone may, in some situations, be insufficient to resolve the keratinocyte immunophenotype. Furthermore, such considerations emphasise the importance of characterising the epidermal responses alongside the immune signals in molecular studies of AD.

In summary, *in-silico* deconvolution of the transcriptional phenotype of AD keratinocytes has revealed two levels of pathology. First, individuals with AD epidermis demonstrate keratinocytes sensing of type 2 cytokines. Secondly, although the IL-4/IL-13 signal becomes enhanced in chronic AD lesions, it appears that induction of an IL-17 response acts as a key switch between acute/chronic AD. This confirms the model of sequential activation of helper T-cell responses across the development and chronicity of cutaneous lesions. Finally, we showed that despite disease resolution with both ciclosporin and dupilumab, ciclosporin treatment rebalances KC17 subpopulation comparable to normal skin but does not modify type 2 cytokine sensing. Whereas, dupilumab therapy reverses the KC2 dominance in lesional AD. Taken together, these observations suggest that whilst type 2 cytokines appear to drive the biology of AD, the efficacy of ciclosporin in AD is likely to lie beyond the targeting of T cells with resultant Th17 inhibition and that other pathways modified by this effective therapy should be explored as potential therapeutic targets.

## Data Availability

All data available through GEO

## Abbreviations

AD: Atopic dermatitis
AD-AcL: Lesional (acute) AD skin
AD-ChL: Lesional (chronic) AD skin
AD-Les: Lesional (acute or chronic) AD skin
AD-Non: Non-lesional AD skin
CD4: Cluster of differentiation 4
CD8: Cluster of differentiation 8
DDC: CD11c^+^ Dermal Dendritic Cell
FDR: False Discovery Rate
GCRMA: Gene Chip Robust Multiarray Averaging
GO: Gene Ontology
GOBP: Gene Ontology, Biological Process
HH: Healthy
IFN: Interferon
IL-1: Interleukin-1
IL-13: Interleukin-13
IL-17: Interleukin-17
IL-22: Interleukin-22
IL-26: Interleukin-26
IL-4: Interleukin-4
IL4Rα: Interleukin-4 Receptor, alpha subunit
KC: Keratinocytes
KC1: Keratinocyte transcriptomic programme to type 1 inflammation
KC17: Keratinocyte transcriptomic programme to type 17 inflammation
KC2: Keratinocyte transcriptomic programme to type 2 inflammation
KC22: Keratinocyte transcriptomic programme to type 22 inflammation
KFG: Keratinocyte Growth Factor
*KRT6A*: Keratin 6A gene
*KRT6B*: Keratin 6B gene
LC: Langerhans Cell
LIMMA: Linear Models for Microarray Data
MCL: Markov Clustering algorithm
Ps-Les: Lesional psoriatic skin
Ps-Non: Non-lesional psoriatic skin
*S100*: S100 protein family gene
SCORAD: SCORing AD index
SCORAD50: 50% improvement in baseline SCORAD index
*SERPIN*: Serine protease inhibitor family gene
Th1: Type 1 inflammation
Th17: Type 17 inflammation
Th2: Type 2 inflammation
Th22: Type 22 inflammation
TNF: Tumour Necrosis Factor

## Acknowledgements

We thank our funding bodies the MRC and Wellcome Trust, and the iCASE PhD studentship sponsor, Unilever, for providing the resources to undertake this research. We also thank Rebecca Ginger, formerly of Unilever (Colworth, UK) for previous discussions and contributions.

## Supporting information

### Supplementary Data 1

#### Co-expression clusters from Figure 1 (50 clusters)

50 gene clusters were identified using co-expression analysis (gene-to-gene expression correlation Pearson *r* >0.7, and MCL 3.1). The gene lists (first tab of Data E1_co-expression genes and GOBP.xlsx file) was input to ToppGene for gene ontology annotation for biological process meaning (GOBP) (ToppGene results tabs labelled 1-50 in Data E1_co-expression genes and GOBP.xlsx file.

See file: Data S1_co-expression genes and GOBP.xlsx

**Supplementary Figure S1.**

**Resolution of whole skin samples into constituent cellular profiles using machine learning**.

**(a)** Deconvolution of keratinocyte signature into profiles representing keratinocyte immunophenotypes across the six skin conditions: healthy (HH), AD non-lesional (AD-Non), AD acute lesional (AD-AcL), AD chronic lesional (AD-ChL), psoriasis non-lesional (Ps-Non), psoriasis lesional (Ps-Les). (Default CIBERSORT settings, training signatures: keratinocytes (unstimulated, stimulated with IFNα, IFNγ, IL-17, IL-1β, TNF, IL-22, IL26, KGF); fibroblasts; melanocytes; CD4+, CD8+ and regulatory T cells; dermal dendritic cells; and, steady-state and activated Langerhans cells here, supplementary table E4) **(b-f)** Individual keratinocyte immunophenotypes; **(b)** unstimulated KC; **(c)** response to IL-1; **(d)** response to keratinocyte growth factor (KGF); **(e)** response to TNF-α; **(f)** response to IL-26 across the five tissue types. Healthy patients (n=14, green), AD non-lesion patients (n=18, red), AD acute lesion patients (n=7, cyan), AD chronic lesion patients (n=18, blue), psoriasis non-lesion patients (n=14, orange), and psoriasis lesion patients (n=14, magenta). ANOVA Sidak’s multiple test bars below: p>0.05,ns; p<0.05, *; p<0.01, **; p<0.001, ***; p<0.0001, ****. Error bars show mean± SD. (Additional to KC fractions shown in main figure 3b-e).

See file: Supplementary figure S1.pdf

**Supplementary Table 1.**
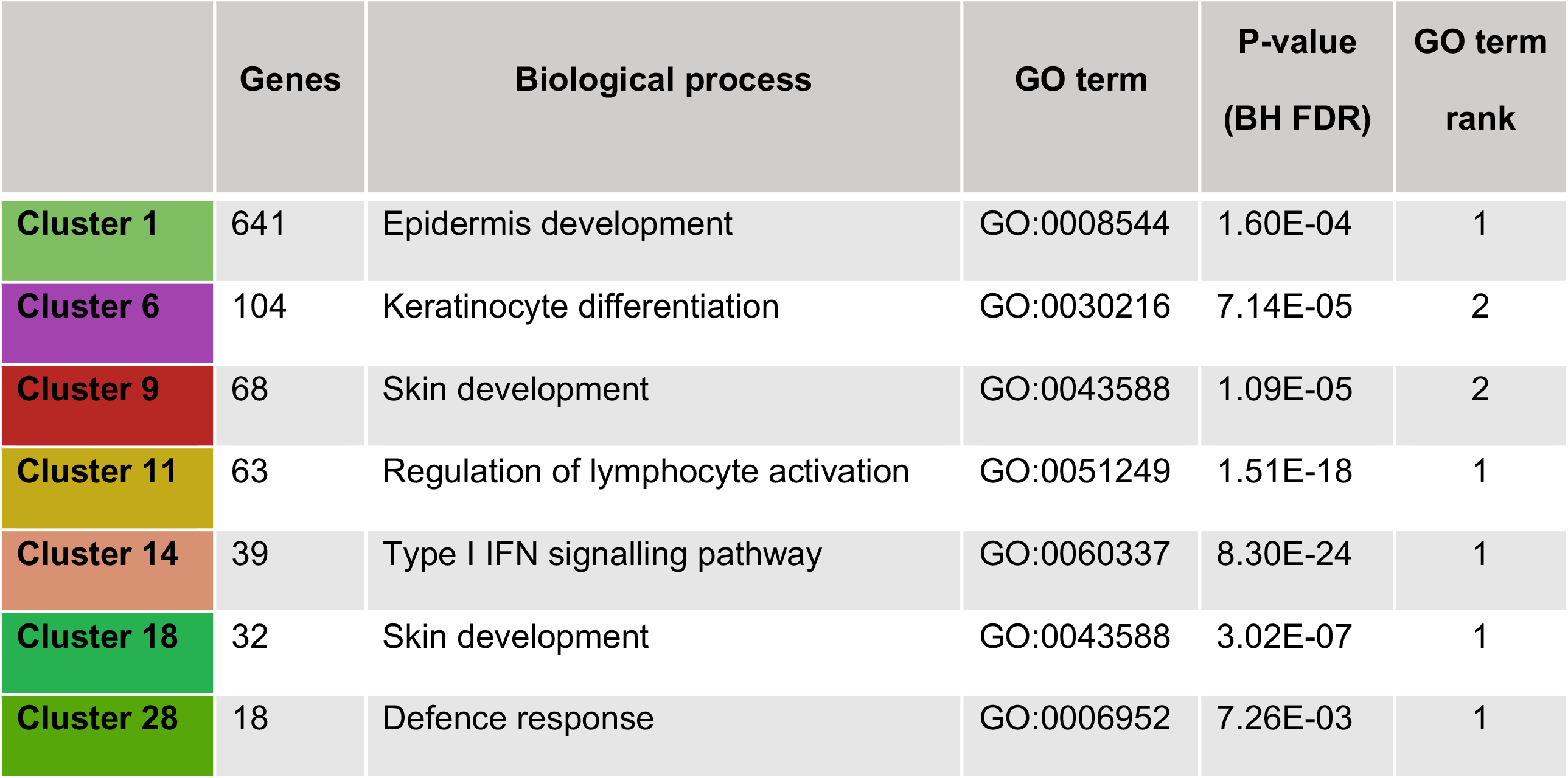
Gene ontology annotation of the cell-based co-expression clusters from whole skin microarray data. Seven cell-based clusters were annotated from gene co-expression analysis (Figure 1A). The clusters were annotated for biological process gene ontology (GOBP) using the ToppFun/ToppGene; the top 200 annotations with FDR Benjamini-Hochberg-corrected p-values <0.05 were selected and collapsed for redundancy using REVIGO. The most significant collapsed annotation from REVIGO was selected as the cluster biological process annotation, with the ToppGene rank shown.

|  | Genes | Biological process | GO term | P-value<br>(BH FDR) | GO term<br>rank |
| --- | --- | --- | --- | --- | --- |
| **Cluster 1** | 641 | Epidermis development | GO:0008544 | 1.60E-04 | 1 |
| **Cluster 6** | 104 | Keratinocyte differentiation | GO:0030216 | 7.14E-05 | 2 |
| **Cluster 9** | 68 | Skin development | GO:0043588 | 1.09E-05 | 2 |
| **Cluster 11** | 63 | Regulation of lymphocyte activation | GO:0051249 | 1.51E-18 | 1 |
| **Cluster 14** | 39 | Type I IFN signalling pathway | GO:0060337 | 8.30E-24 | 1 |
| **Cluster 18** | 32 | Skin development | GO:0043588 | 3.02E-07 | 1 |
| **Cluster 28** | 18 | Defence response | GO:0006952 | 7.26E-03 | 1 |

**Supplementary Table 2**.

Training samples of unstimulated keratinocytes (GSE36287) and skin dermal fibroblasts (GSE34308) with expression reduced to mean expression across replicates.

See file: Table S2.xlsx

**Supplementary Table 3**.

Training samples of unstimulated keratinocytes (GSE36287); skin dermal fibroblasts (GSE34308); CD4+, CD8+ and regulatory T cells (GSE74158), melanocytes (GSE4570), activated Langerhans cells and CD11C+ dermal dendritic cells (GSE49475); and steady state Langerhans cells (GSE23618), with expression reduced to mean expression across replicates.

See file: Table S3.xlsx

**Supplementary Table 4**.

Training samples of keratinocytes (unstimulated, stimulated with IFNα, IFNγ, IL-17, IL-1β, TNF, IL-22, IL26, KGF (GSE36287, GSE7216)); skin dermal fibroblasts (GSE34308); CD4+, CD8+ and regulatory T cells (GSE74158), melanocytes (GSE4570), activated Langerhans cells and CD11C+ dermal dendritic cells (GSE49475); and steady state Langerhans cells (GSE23618), with expression reduced to mean expression across replicates.

See file: Table S4.xlsx

## References

1. Biedermann T, Skabytska Y, Kaesler S, Volz T. Regulation of T Cell Immunity in Atopic Dermatitis by Microbes: The Yin and Yang of Cutaneous Inflammation. Front Immunol. 2015;6:353.

2. Ungar B, Garcet S, Gonzalez J, Dhingra N, Correa da Rosa J, Shemer A, et al. An Integrated Model of Atopic Dermatitis Biomarkers Highlights the Systemic Nature of the Disease. J Invest Dermatol. 2017;137:603–13.

3. Suárez-Fariñas M, Tintle SJ, Shemer A, Chiricozzi A, Nograles K, Cardinale I, et al. Nonlesional atopic dermatitis skin is characterized by broad terminal differentiation defects and variable immune abnormalities. J Allergy Clin Immunol. 2011;127:954-64.e1-4.

4. Guttman-Yassky E, Bissonnette R, Ungar B, Suárez-Fariñas M, Ardeleanu M, Esaki H, et al. Dupilumab progressively improves systemic and cutaneous abnormalities in patients with atopic dermatitis. J Allergy Clin Immunol. 2019;143:155–72.

5. Gittler JK, Shemer A, Suárez-Fariñas M, Fuentes-Duculan J, Gulewicz KJ, Wang CQF, et al. Progressive activation of T(H)2/T(H)22 cytokines and selective epidermal proteins characterizes acute and chronic atopic dermatitis. J Allergy Clin Immunol. 2012;130:1344–54.

6. Mansouri Y, Guttman-Yassky E. Immune Pathways in Atopic Dermatitis, and Definition of Biomarkers through Broad and Targeted Therapeutics. J Clin Med. 2015;4:858–73.

7. Ariëns L, Gadkari A, van Os-Medendorp H, Ayyagari R, Terasawa E, Kuznik A, et al. Dupilumab versus Cyclosporine for Treatment of Moderate-to-Severe Atopic Dermatitis in Adults: Indirect Comparison Using the Eczema Area and Severity Index. Acta Derm Venereol. 2019;0.

8. Brunner PM, Guttman-Yassky E, Leung DYM. The immunology of atopic dermatitis and its reversibility with broad-spectrum and targeted therapies. J Allergy Clin Immunol. 2017;139:S65–76.

9. Hönzke S, Wallmeyer L, Ostrowski A, Radbruch M, Mundhenk L, Schäfer- Korting M, et al. Influence of Th2 Cytokines on the Cornified Envelope, Tight Junction Proteins, and ß-Defensins in Filaggrin-Deficient Skin Equivalents. J Invest Dermatol. 2016;136:631–9.

10. Nestle FO, Di Meglio P, Qin J-Z, Nickoloff BJ. Skin immune sentinels in health and disease. Nat Rev Immunol. 2009;9:679–91.

11. Pfalzgraff A, Brandenburg K, Weindl G. Antimicrobial Peptides and Their Therapeutic Potential for Bacterial Skin Infections and Wounds. Front Pharmacol. 2018;9:281.

12. Colombo I, Sangiovanni E, Maggio R, Mattozzi C, Zava S, Corbett Y, et al. HaCaT Cells as a Reliable In Vitro Differentiation Model to Dissect the Inflammatory/Repair Response of Human Keratinocytes. Mediators Inflamm. 2017;2017:1–12.

13. Nakagawa S, Matsumoto M, Katayama Y, Oguma R, Wakabayashi S, Nygaard T, et al. Staphylococcus aureus Virulent PSMα Peptides Induce Keratinocyte Alarmin Release to Orchestrate IL-17-Dependent Skin Inflammation. Cell Host Microbe. 2017;22:667-677.e5.

14. Lai Y, Gallo RL. AMPed up immunity: how antimicrobial peptides have multiple roles in immune defense. Trends Immunol. 2009;30:131–41.

15. Selsted ME, Ouellette AJ. Mammalian defensins in the antimicrobial immune response. Nat Immunol. 2005;6:551–7.

16. Nickoloff BJ, Turka LA. Immunological functions of non-professional antigen- presenting cells: new insights from studies of T-cell interactions with keratinocytes. Immunol Today. 1994;15:464–9.

17. Valeri M, Raffatellu M. Cytokines IL-17 and IL-22 in the host response to infection. Napier B, editor. Pathog Dis. 2016;74:ftw111.

18. Archer NK, Adappa ND, Palmer JN, Cohen NA, Harro JM, Lee SK, et al. Interleukin-17A (IL-17A) and IL-17F Are Critical for Antimicrobial Peptide Production and Clearance of Staphylococcus aureus Nasal Colonization. Freitag NE, editor. Infect Immun. 2016;84:3575–83.

19. Dixon BREA, Radin JN, Piazuelo MB, Contreras DC, Algood HMS. IL-17a and IL-22 Induce Expression of Antimicrobials in Gastrointestinal Epithelial Cells and May Contribute to Epithelial Cell Defense against Helicobacter pylori. Ho PL, editor. PLoS One. 2016;11:e0148514.

20. Yano S, Banno T, Walsh R, Blumenberg M. Transcriptional responses of human epidermal keratinocytes to cytokine interleukin-1. J Cell Physiol. 2008;214:1–13.

21. Sa SM, Valdez PA, Wu J, Jung K, Zhong F, Hall L, et al. The effects of IL-20 subfamily cytokines on reconstituted human epidermis suggest potential roles in cutaneous innate defense and pathogenic adaptive immunity in psoriasis. J Immunol. 2007;178:2229–40.

22. Guilloteau K, Paris I, Pedretti N, Boniface K, Juchaux F, Huguier V, et al. Skin Inflammation Induced by the Synergistic Action of IL-17A, IL-22, Oncostatin M, IL-1{alpha}, and TNF-{alpha} Recapitulates Some Features of Psoriasis. J Immunol. 2010;184:5263–70.

23. Guttman-Yassky E, Lowes MA, Fuentes-Duculan J, Zaba LC, Cardinale I, Nograles KE, et al. Low expression of the IL-23/Th17 pathway in atopic dermatitis compared to psoriasis. J Immunol. 2008;181:7420–7.

24. Guttman-Yassky E, Krueger JG. Atopic dermatitis and psoriasis: two different immune diseases or one spectrum? Curr Opin Immunol. 2017;48:68–73.

25. Freeman TC, Goldovsky L, Brosch M, van Dongen S, Mazière P, Grocock RJ, et al. Construction, visualisation, and clustering of transcription networks from microarray expression data. PLoS Comput Biol. 2007;3:2032–42.

26. Theocharidis A, van Dongen S, Enright AJ, Freeman TC. Network visualization and analysis of gene expression data using BioLayout Express(3D). Nat Protoc. 2009;4:1535–50.

27. Chen J, Bardes EE, Aronow BJ, Jegga AG. ToppGene Suite for gene list enrichment analysis and candidate gene prioritization. Nucleic Acids Res. 2009;37:W305–11.

28. Newman AM, Liu CL, Green MR, Gentles AJ, Feng W, Xu Y, et al. Robust enumeration of cell subsets from tissue expression profiles. Nat Methods. 2015;12:453–7.

29. Bigler J, Rand HA, Kerkof K, Timour M, Russell CB. Cross-study homogeneity of psoriasis gene expression in skin across a large expression range. Brandner JM, editor. PLoS One. 2013;8:e52242.

30. Brunner PM, Emerson RO, Tipton C, Garcet S, Khattri S, Coats I, et al. Nonlesional atopic dermatitis skin shares similar T-cell clones with lesional tissues. Allergy. 2017;72:2017–25.

31. Tang TS, Bieber T, Williams HC. Are the concepts of induction of remission and treatment of subclinical inflammation in atopic dermatitis clinically useful? J Allergy Clin Immunol. 2014;133:1615-1625.e1.

32. Lessard JC, Piña-Paz S, Rotty JD, Hickerson RP, Kaspar RL, Balmain A, et al. Keratin 16 regulates innate immunity in response to epidermal barrier breach. Proc Natl Acad Sci U S A. 2013;110:19537–42.

33. Rorke EA, Adhikary G, Young CA, Rice RH, Elias PM, Crumrine D, et al. Structural and biochemical changes underlying a keratoderma-like phenotype in mice lacking suprabasal AP1 transcription factor function. Cell Death Dis. 2015;6:e1647.

34. Chen B, Khodadoust MS, Liu CL, Newman AM, Alizadeh AA. Profiling Tumor Infiltrating Immune Cells with CIBERSORT. Methods Mol Biol. 2018;1711:243–59.

35. Esaki H, Ewald DA, Ungar B, Rozenblit M, Zheng X, Xu H, et al. Identification of novel immune and barrier genes in atopic dermatitis by means of laser capture microdissection. J Allergy Clin Immunol. 2015;135:153–63.

